# Audit of Intraoperative Hypertension Management in 75 Patients at THQ Hospital Sadiq Abad

**DOI:** 10.1101/2025.10.14.25337963

**Authors:** Muhammad Mubashar Javed

**Affiliations:** THQ Hospital Sadiq Abad

**Author notes:** Correspondence |.

## Abstract

**Background:** Intraoperative hypertension (IOH) is a frequent anesthetic challenge linked to myocardial ischemia, arrhythmias, cerebrovascular events, and increased surgical bleeding. Despite its impact, structured evaluation of IOH management is inconsistent, especially in resource-limited hospitals.

**Objective:** To evaluate recognition, documentation, precipitating causes, and management of IOH at THQ Hospital Sadiq Abad, and to identify gaps relative to accepted standards.

**Methods:** A retrospective audit of 75 adult patients (18–75 years) with documented IOH episodes from January to June 2025 was conducted. Data extracted from anesthesia records included demographics, ASA grade, type of surgery, anesthetic technique, recorded trigger(s), interventions, and immediate outcomes. Audit standards were adapted from the Association of Anaesthetists’ peri-operative hypertension guidance. Descriptive statistics were used to summarize findings and guide recommendations.

**Results:** IOH was documented in all 75 cases (100%). A precipitating cause was recorded in 52%. The most common first-line response was deepening anesthesia (42%), followed by opioid boluses (28%) and antihypertensives (16%). Targeted correction of reversible triggers (e.g., bladder decompression, ventilatory adjustment) was documented in 14%. No peri-operative mortality occurred; however, prolonged IOH (>15 minutes) was noted in a minority of cases.

**Conclusion:** While recognition of IOH was universal, documentation of triggers and targeted, protocolized management were suboptimal. Introducing a structured proforma, theatre-posted algorithms, and focused teaching—followed by re- audit in 6–12 months—may improve patient safety.

## Introduction

Intraoperative hypertension (IOH)—commonly defined as a sustained increase in systolic blood pressure or mean arterial pressure exceeding 20% above baseline—occurs across a wide spectrum of procedures. Pathophysiology is multifactorial and includes inadequate depth of anesthesia, insufficient analgesia, sympathetic surges during surgical stimulation, hypercarbia, hypoxia, and reversible factors such as urinary bladder distension. If unrecognized or undertreated, IOH increases myocardial oxygen demand, may precipitate arrhythmias, exacerbate bleeding, and potentially worsen neurologic outcomes.

Although perioperative hypotension has historically received greater attention, contemporary literature underscores that hypertensive excursions are also independently associated with adverse events. In low- and middleincome settings, additional challenges—limited surveillance, constrained drug formularies, and variable documentation—can compound the risk. Yet there is sparse published experience from district-level hospitals in Pakistan.

Clinical audit—measuring practice against standards, implementing change, and reauditing—is a pragmatic strategy for quality improvement. This audit at THQ Hospital Sadiq Abad aimed to characterize how IOH is recognized and managed locally, to benchmark current performance against accepted standards, and to generate practical, context-aware recommendations.

## Methods

Study design and setting: A retrospective clinical audit was conducted in the operating theatres of THQ Hospital Sadiq Abad from January to June 2025. The theatres serve general surgery, gynecology, orthopedics, ENT, and minor procedures.

Inclusion and exclusion: Adult patients (18–75 years) with a documented IOH episode were eligible. Exclusions included ASA IV–V, pediatric cases, and incomplete or illegible records.

Standards and data items: Audit standards were adapted from the Association of Anaesthetists perioperative hypertension guidance and standard anesthesia texts (Miller; Morgan & Mikhail). Key criteria were: (1) IOH episode documented; (2) likely precipitating cause recorded; (3) targeted correction attempted; (4) timely use of antihypertensives for persistent or severe IOH; (5) postoperative note of hemodynamic issues. Data fields included demographics, ASA grade, surgery type, anesthetic technique, triggers, interventions, and outcomes.

Analysis and governance: Descriptive statistics were performed (frequencies, percentages, crosstabulations). As an audit of anonymized routine records, formal ethical approval was not required; institutional permission was obtained.

## Results

Seventy-five patients met inclusion criteria. Mean age was 42 ± 15 years; 64% were male. Most patients were ASA I–II (83%). IOH occurred across all specialties, with general surgery constituting 40% of cases. A precipitating cause was documented in 52%: inadequate analgesia (≈35%), light anesthesia (≈28%), bladder distension (≈12%), hypercarbia (≈7%), and unclassified (≈18%).

Management patterns showed a preference for deepening anesthesia (42%) and opioid supplementation (28%), with antihypertensive agents used in 16%. Targeted correction of reversible triggers was noted in 14%. No peri-operative mortality occurred; several cases exhibited prolonged IOH (>15 minutes), highlighting the importance of standardized responses.

**Table 1.**
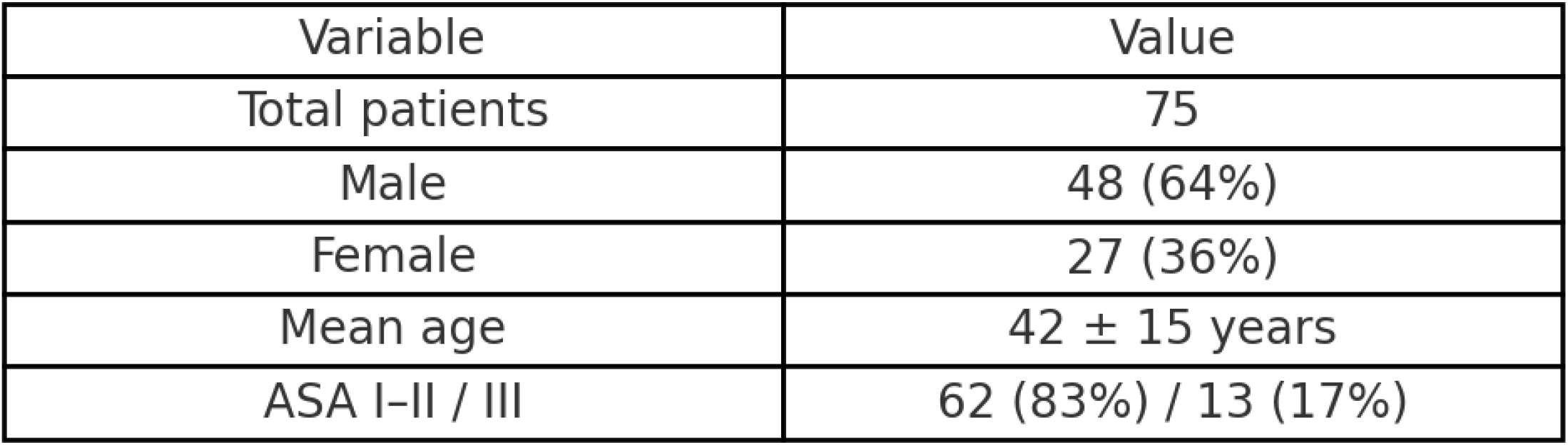
Demographic characteristics of patients.

**Table 2.** Distribution of surgical categories.

| Category | n (%) |
| --- | --- |
| General Surgery | 30 (40%) |
| Gynecology | 15 (20%) |
| Orthopedics | 13 (18%) |
| ENT | 7 (10%) |
| Other | 10 (12%) |

**Figure 1.**
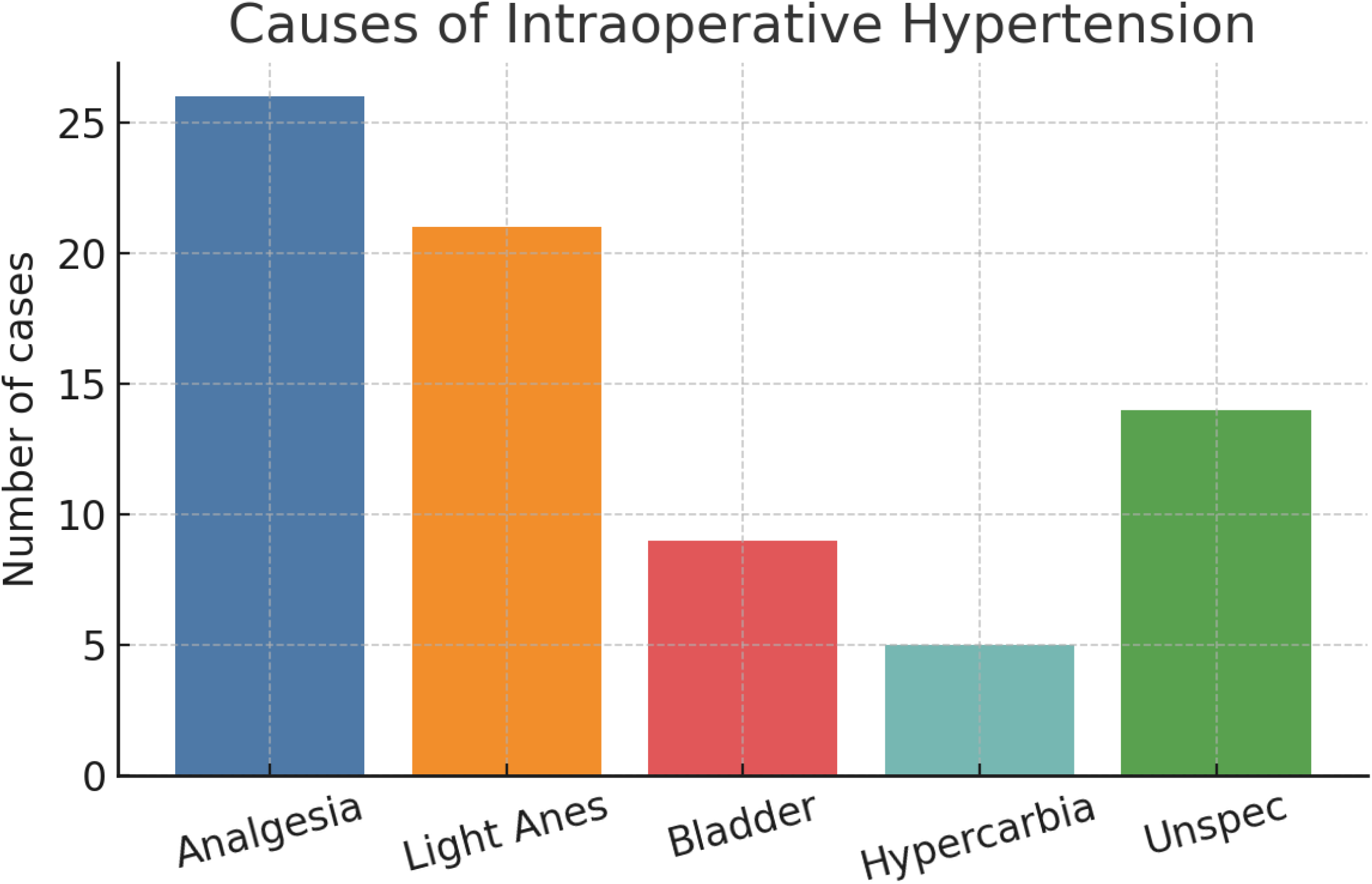
Causes of intraoperative hypertension.

**Figure 2.**
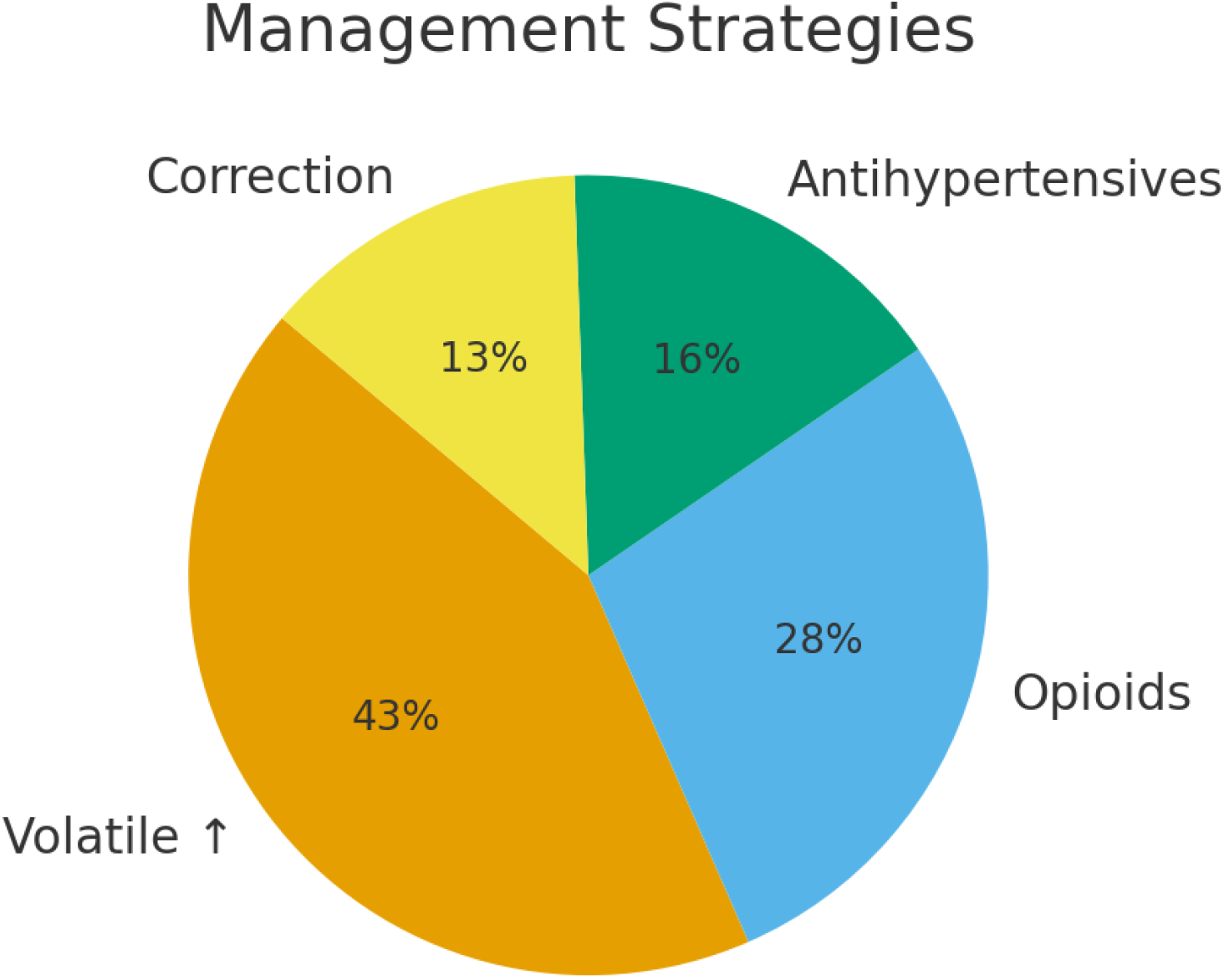
Management strategies used.

## Discussion

This audit demonstrates universal recognition and documentation of IOH episodes but suboptimal documentation of precipitating causes and relatively low utilization of antihypertensive drugs compared to international reports. The dominant reflex— deepening anesthesia—can be efficacious yet risks masking reversible triggers such as hypercarbia or bladder distension. A structured approach that first verifies the reading (correct cuff size, repeat NIBP/consider IBP), then rapidly screens for common triggers, and finally escalates to titrated antihypertensives in persistent or high-risk scenarios, is likely to reduce exposure time to hypertensive stress.

Strengths of this work include consecutive case capture, explicit standards, and actionable recommendations contextualized to a district-level hospital. Limitations include a retrospective design, single-center scope, and reliance on existing documentation. Future work could incorporate prospective data collection, process measures (time-to-intervention), and patient-centered outcomes (troponin, AKI, 30-day complications).

## Recommendations

1. Implement a one-page intraoperative hypertension proforma with prompts for likely triggers and specific actions.
2. Post the management algorithm (Figure 3) in all theatres and include it in trainee orientation.
3. Ensure availability of first-line antihypertensives (labetalol, hydralazine, esmolol) and a GTN infusion protocol.
4. Establish a three-month IOH logbook to monitor adherence, response times, and outcomes.
5. Conduct a targeted teaching session and simulation drills on IOH management.
6. Reaudit in 6–12 months to evaluate improvements in documentation and targeted management.

**Figure 3.**
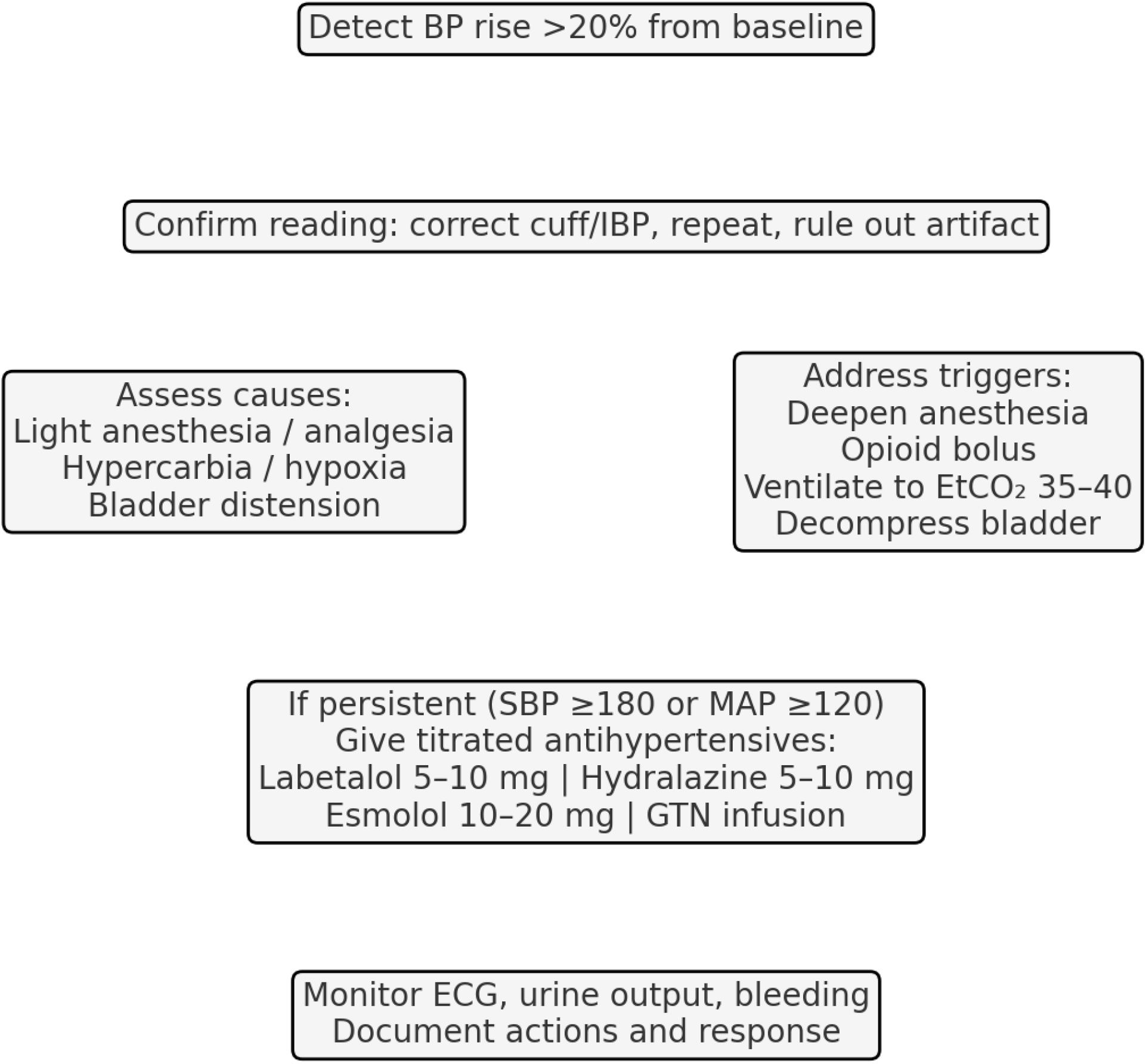
Stepwise intraoperative hypertension management algorithm.

**Figure 4.**
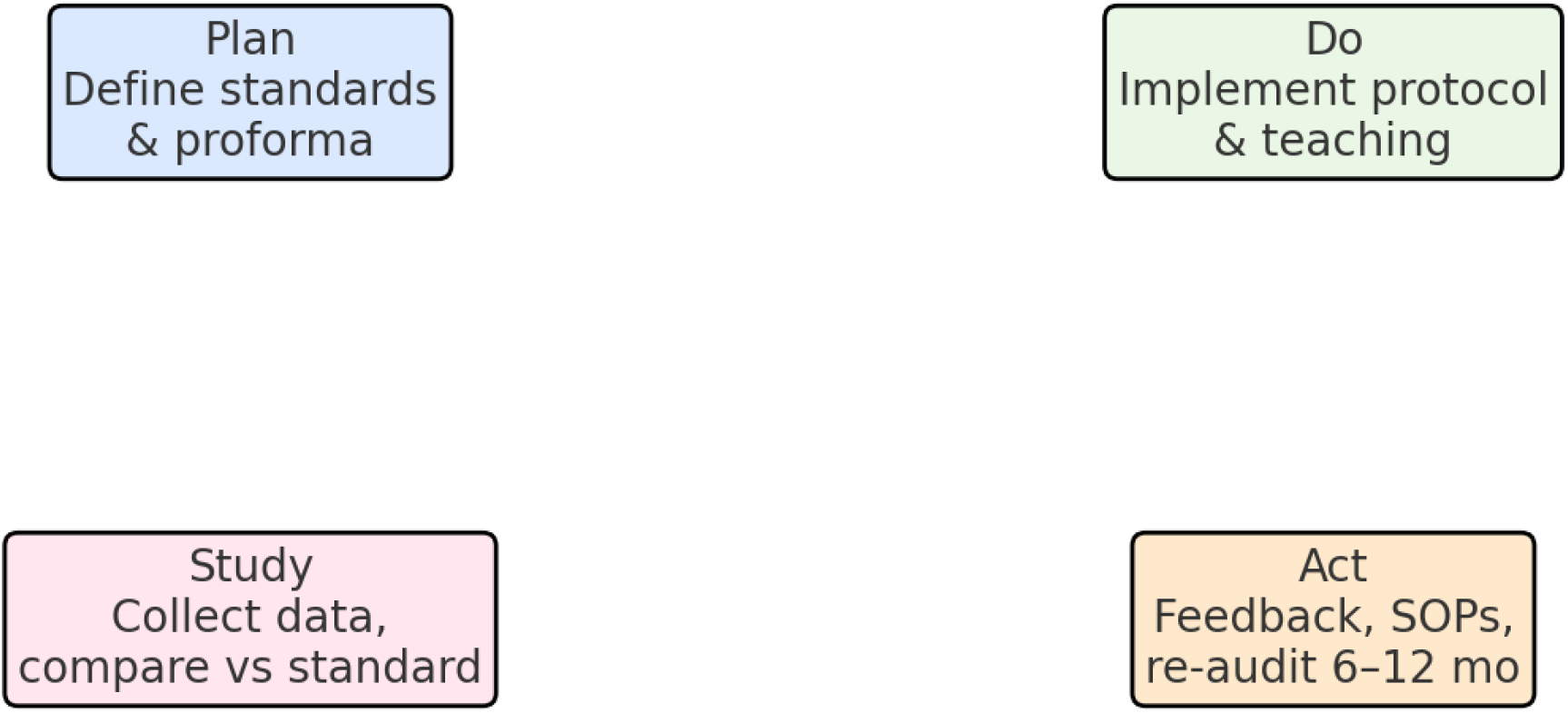
Audit cycle (Plan–Do–Study–Act) for sustained improvement.

**Figure 5.**
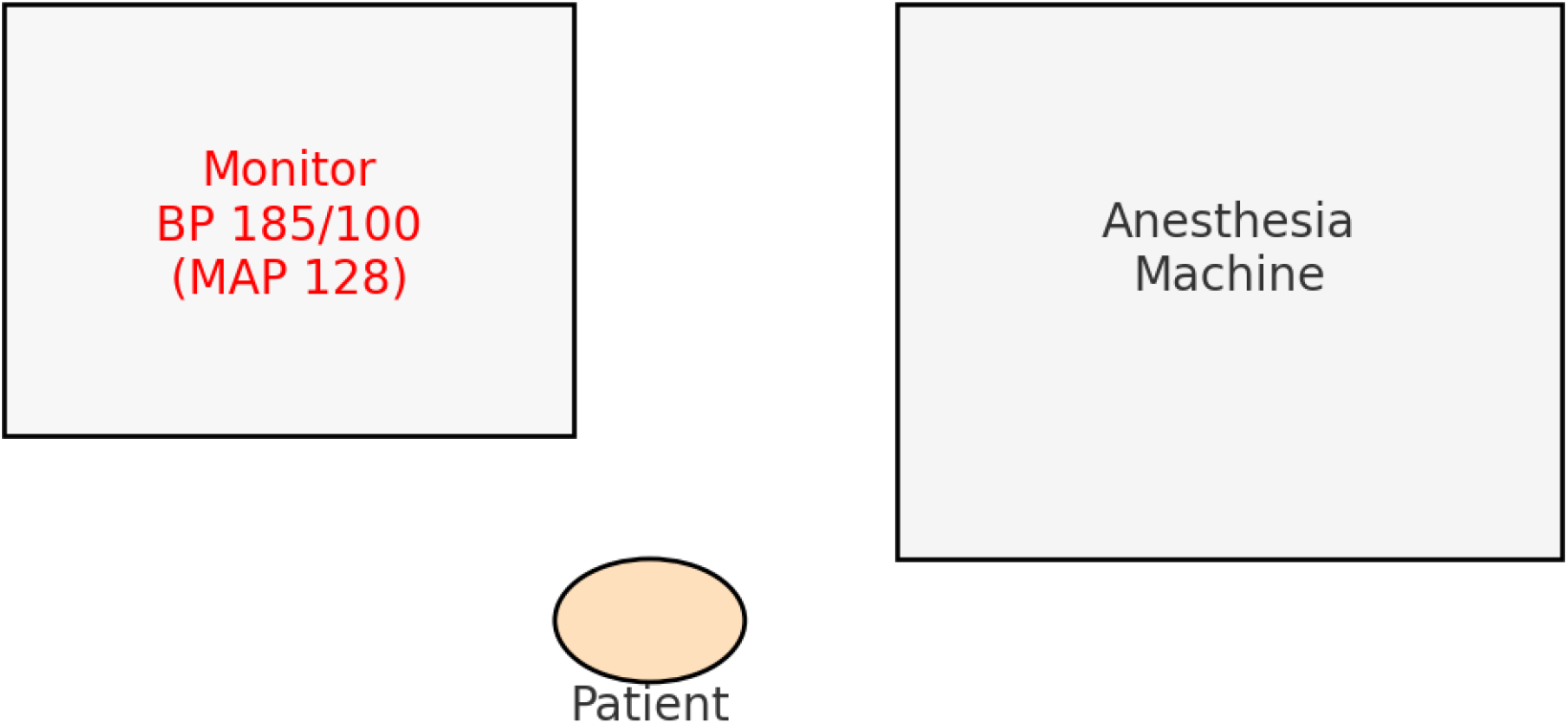
Operating room schematic: monitoring and anesthesia station.

## Data Availability

Data available upon reasonable request

